# Classifying Drug Ratings Using User Reviews with Transformer-Based Language Models

**DOI:** 10.1101/2021.04.15.21255573

**Authors:** Akhil Shiju, Zhe He

## Abstract

Drugs.com provides users’ textual reviews and numeric ratings of drugs. However, text reviews may not always be consistent with the numeric ratings. Overly positive or negative rating may be misleading. In this project, to classify user ratings of drugs with their textual reviews, we built classification models using traditional machine learning and deep learning approaches. Machine learning models including Random Forest and Naive Bayesian classifiers were built using TF-IDF features as input. Also, transformer-based neural network models including BERT, BioBERT, RoBERTa, XLNet, ELECTRA, and ALBERT were built using the raw text as input. Overall, BioBERT model outperformed the other models with an overall accuracy of 87%. We further identified UMLS concepts from the postings and analyzed their semantic types in the postings stratified by the classification result. This research demonstrated that transformer-based models can be used to classify drug reviews and identify reviews that are inconsistent with the ratings.

## Introduction

The evaluation of the efficacy and safety of drugs heavily relies on randomized controlled trials with rigorous inclusion and exclusion criteria.^1^ However, such processes are limited to a small number of individuals enrolled in the study and are constrained to participants in the target population who meet possibly restrictive eligibility criteria, limiting the population representativeness and subsequent study generalizability.^2,3^ The ramifications of these acclimations could potentially have resulted in the overestimation of the efficacy of the product and misidentification of adverse events/side effects in the diverse population.^4^ To counter such issues, approaches such as post-marketing drug surveillance have been introduced to optimize the safety of the drug after its regulatory approval and mass production.^5^

There are two major forms of post-marketing drug surveillance. Some are formed by government regulators such as the Vaccine Adverse Event Reporting System (VAERS) by the United States Food and Drug Administration^6^ or the Yellow Card Scheme by the United Kingdom Medicines and Healthcare Products Regulatory Agency.^7^ Also, public/private organizations have a system to monitor drug side-effects such as the Research on Adverse Drug events And Reports.^8^ Existing methods for identifying adverse events typically focused on analyzing molecular drug composition,^9^ query logs,^10^ VAERS records,^11^ or clinical notes in the medical records^12^ but did not analyze specifically the sentiment of the consumers using their reviews of the drug.^13^ The application of post-market drug surveillance has been successfully applied in the identification of adverse events through safety reports by the introduction of deep learning-based methods including the extraction of temporal events, the procedure performed, and social circumstance^14^.

In the era of Web 2.0, the Internet has opened up new pathways to obtain information directly from consumers about their drug reviews in an elaborative format. Publicly available information on the Internet offers an easily attainable resource that could be leveraged to gain a deep understanding of the drug reviews by the users. Entire user reviews are fully available on drug review websites, on which users can comment on their personal experiences of the drugs they have taken for a specific condition. Unlike many other forms of medical data, this information is not filtered through medical professionals. Since these reviews are given by anonymous users, there is no risk of patient health record violation for confidentiality. These reviews contain a plethora of information regarding individual experiences associated with the drugs such as symptoms, adverse events, and interactions with other drugs. Such reviews have also contained an extensive amount of user sentiment related to a particular condition, which could be leveraged to detect the side effects and efficacy of drugs.^15^

However, many barriers exist in the extraction of sentiment from these online medical reviews. For instance, user reviews of drugs in such online forms are typically unconventional and most reviewers lack medical knowledge, posing barriers for extracting meaningful information from them. In addition, many review websites use some form of numerical rating that has served the role of quantifying such a sentiment, but they do not provide a clear guideline for giving a certain numeric rating. As such, these review websites may have introduced biases as individual users may have different perception as to what a high score means versus what would have constituted a low score. Users have tended to reduce the effort required in reporting values by rating all qualities as highly important, thus resulting in overly positive ratings.^16^ This could lead to an unintended positive view of the overrated drugs by the general public, albeit less effective for certain population subgroups. Prior research has found that web-based reviews have the potential to be viewed as an applicable source of information for analysis, but the direct reliance on consumer ratings could be biased by the consumer experience. For example, addictive drugs have been observed to be typically highly rated in comparison to other drugs which have treated the same condition, even if these additive drugs underperformed.^17^ Thus, the ratings of those drugs may be skewed, thus a potential solution could be analyzing the relationship between the review and the rating and identify skewed ratings based on the textual review.^18^

The application of machine learning, especially through transformer-based language models pre-trained with an enormous amount of data, offers a unique approach to classify textual information.^19^ In this project, we evaluated the feasibility of leveraging machine learning and natural language processing to classify user ratings based on their textual review to identify the locations of contingency. In addition, the constructed models can then be tested to identify overly positive and overly negative instances. To provide some interpretability of the classification results, we used an interpretation tool called Eli5 to highlight phrases in the text that have a positive or negative impact on the classification results. The overly positive (false negative) or overly negative (false positive) scores (user rating that was incorrectly classified by the model) were further analyzed with QuickUMLS to identify semantic type patterns associated with these classifications.

## Methods

### Dataset

We obtained the dataset from the UCI Machine Learning Repository.^20^ These instances were collected from Drugs.com using Beautiful Soup. The dataset used for this study consists of user drug reviews, drug names, related medical conditions, and a 10-point rating. The rating were integer values ranging from 1 to 10 with 10 being the highest possible rating. Table 1 shows example records of the dataset. Figure 1 shows the distribution of reviews by ratings. The ratings were shown to be skewed to the left to suggest that most drugs received a relatively high score. Prior analysis of this dataset focused primarily on the sentiment analysis^21^ and classification of reviews used an n-gram technique which used unequal classes, thus skewing accuracy^22^. Neither was there an emphasis on the error analysis of the models. In total, the dataset consists of 215,063 instances. The numeric ratings had a mean of 7.00 with a standard deviation of 3.27. There are 836 classified medical conditions in the dataset.

**Table 1.** Two examples of a high-rating review versus a low-rating review with condition, drug name, and rating.

| Drug Name | Condition | Review | Rating |
| --- | --- | --- | --- |
| Chantix | Smoking Cessation | I smoked for 50+ years. Took it for one week and that was it. I didn't think it was possible for me to quit. It has been 6 years now. Great product. | 10.0 |
| Excedrin | Migraine | Does not work for people sensitive to caffeine. I was jittery and nervous and queasy after using a single dose. | 2.0 |

**Figure 1.**
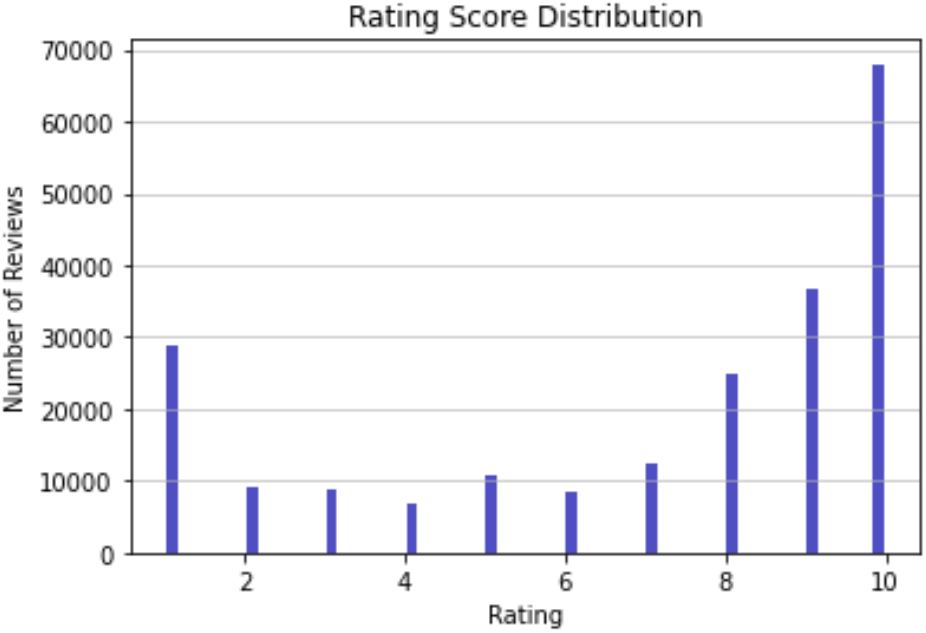
Total number of reviews in the dataset.

### Review Rating Classification

Since the primary focus of this study was to classify textual reviews, the data was broken down using a median of the ratings: ratings 8 or above were considered *above average*, and below 8 were considered as *below average*. The binary classification was chosen over multiple classes since the other objective of this project was to identify overly positive and overly negative ratings which could be identified by misclassification of the system versus the actual score. Thus, misclassification identification in its simplest sense would only be possible with a binary system. Instances in which the reviews contained more than 514 tokens were removed from the study due to the input size limit of the transformer-based language models.

The common methodology for transfer learning has been applied through the application of pre-training on a large unannotated corpus that was capable of understanding the composition of the data type such as patterns in the language. This process could be considered as self-supervised learning. This pre-trained model is then followed by the fine-tuning process which focused on the training on an application-specific dataset.

BERT: Some common language models are pre-trained by predicting the next word in a sequence, but Bidirectional encoder representation from transformer (BERT) looked at bidirectional predicting context masked intermediate text tokens in the pretraining from Wikipedia and BookCorpus and next sentence prediction. Bert-base-uncased was used for this project^23^.

BioBERT: The BERT model has been pre-trained with a medical corpus from publicly available data from PubMed and PMC.^24^ The model which was used was from Huggingface labeled Bio_ClinicalBERT.

ALBERT: A Lite BERT (ALBERT) is a model which focused on being a less memory-heavy and faster version of BERT through the separation of the word embedding into two matrixes and by cross-layer parameter sharing.^25^ Albert-base-v2 was used for this model.

RoBERTa: Robustly Optimized BERT Approach (RoBERTa) has been considered a pretraining model that eliminates the next sentence prediction task and adapts a novel approach of dynamic masking which randomized the masked token between training epochs.^26^ RoBERTa outperformed BERT of multiple results such as GLUE, RACE, and SQuAD. Roberta-base was the model selected for this project.

XLNet: As a more computationally expensive model, the Generalized Auto-Regressive model (XLNet) implemented a system where it applies an autoencoder language model.^27^

ELECTRA: Efficiently Learning an Encoder that Classifies Token Replacements Accurately (ELECTRA) replaced the masked language task with a generator and pre-trains the model to identify which token has been replaced.^28^ The Electra-base-discriminator was used for this project.

We split the dataset into a training set (60%), a validation set (20%), and the test set (20%). These datasets were further classified into lists which were then converted into Transformer datasets that could be trained by a neural network to generate a model.

We constructed these transformer-based text classification models utilizing the Huggingface transformers using the Python k-train pipeline wrapper class for text classification. The models used for this project consisted of BioBERT, ELECTRA, RoBERTa, XLNet, ALBERT, and BERT. The parameter included a 514 max token length, a 5e^-5 learning rate, and a batch size of 6. The train test dataset was fed into the neural network trained to minimize validation data loss. After the training was completed, a confusion matrix of the test data was generated to determine F1 scores for the classes and the accuracy in comparison to the user ratings.

As a baseline approach for evaluating the transformer-based models, bag-of-words (BOW) models were constructed based on term frequency and inverse document frequency (TF-IDF). The textual reviews were converted into a bag of words representation. Afterward, a term TF-IDF score matrix was computed for the bag of words representation. We trained and evaluated a Random Forest classifier and a Naïve-Bayes classifier with the BOW features.

The test data was stratified for the top 10 conditions based on the test data user reviews as seen in Table 2. The transformer models were then used to classify each of the different conditions to determine condition-specific F1 score and accuracy. The overall workflow is outlined in Figure 2.

**Table 2.**
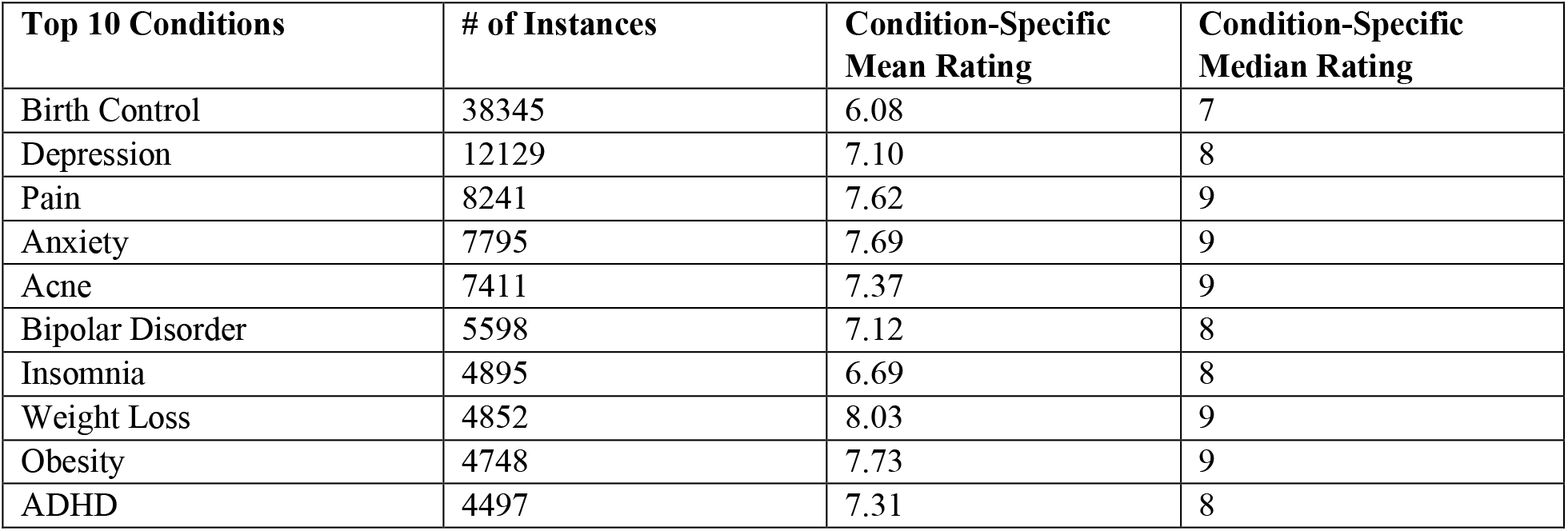
Condition-specific statistics. Birth Control was the most common condition which users reviewed followed by depression, pain, anxiety, and acne.

**Figure 2.**
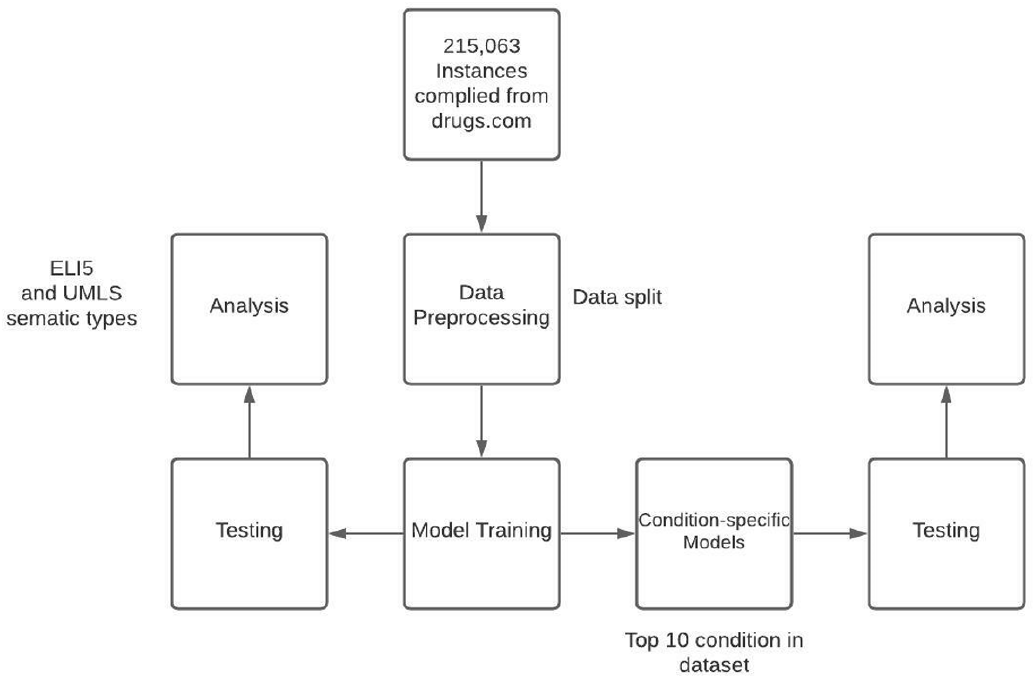
The workflow of the project.

### Model Interpretation

After the best-performing transformer model was selected, to provide some interpretability for the model, Eli5 metrics were applied to the model. Eli5 has been used to understand why a certain classification through the identification of important features such as highlighting significant text features.^29^ This is accomplished by inspecting the model parameters to discover the global implications. This was performed for many reviews to establish some sense of how the model performed these classifications. Top scores were also computed through the Eli5 metrics.

### Error Analysis

An analysis of the potential relationship between false positives, false negatives, true positives, true negatives from the best overall performing models was conducted by analyzing the occurrences of certain semantic types of the Unified Medical Language System (UMLS) Metathesaurus, which links terms to biomedical concepts.^30^ We would like to see whether certain error types had deviation in the semantic types present in the review in comparison to the other conditional cases. This was conducted using the QuickUMLS package, an unsupervised tool for biomedical term extraction using simstring.^31^ We chose 8 semantic types that were most prevalent in the dataset and had some medical significance, including Sign or Symptom, Disease or Syndrome, Organism Function, Pathologic Function, Body Substance, Body Location, or Region, Body Part, Organ, or Organ Component, and Health Care Activity. Only instances with a 1.0 Jaccard similarity were retained, and the best matching CUIs were selected. After the semantic types were extracted for all the reviews, the means were calculated by true positive, true negative, false positive and false negative (class type). A one-way ANOVA was employed to determine whether there was a significant difference based on the mean value of the number of concepts of a certain semantic type per post across different class types.

## Results

### Classification of the Drug Review Rating

Overall, the model generated by the BioBERT and ELECTRA outperformed the other models on a variety of metrics as displayed in Table 3. The BOW models showed lower accuracy compared to the other constructions. XLNet had the longest training time compared to the other models. Table 4 provides the condition-specific statistics for the top 10 conditions. The ratings of the reviews pertaining to the Birth Control, Depression, Pain drugs were classified with high accuracy than the ratings of the drugs for other conditions. The conditions with lower instances had lower accuracy than the conditions with higher instances. However, there are many notable deviations present such as the pain and obesity models’ lower accuracy or the higher accuracy for the ADHD model.

**Table 3.** Overall condition validation from the test dataset for the minimized loss for the top-performing models.

| Model | Above Average F1 | Below Average F1 | Accuracy |
| --- | --- | --- | --- |
| BERT | 0.84 | 0.84 | 0.84 |
| RoBERTa | 0.83 | 0.83 | 0.84 |
| XLNet | 0.84 | 0.84 | 0.84 |
| BioBERT | **0.87** | **0.87** | **0.87** |
| ELECTRA | 0.85 | 0.87 | 0.86 |
| ALBERT | 0.75 | 0.81 | 0.78 |
| Random Forest (BOW) | 0.77 | 0.45 | 0.68 |
| Naïve Bayes (BOW) | 0.76 | 0.03 | 0.61 |

**Table 4.**
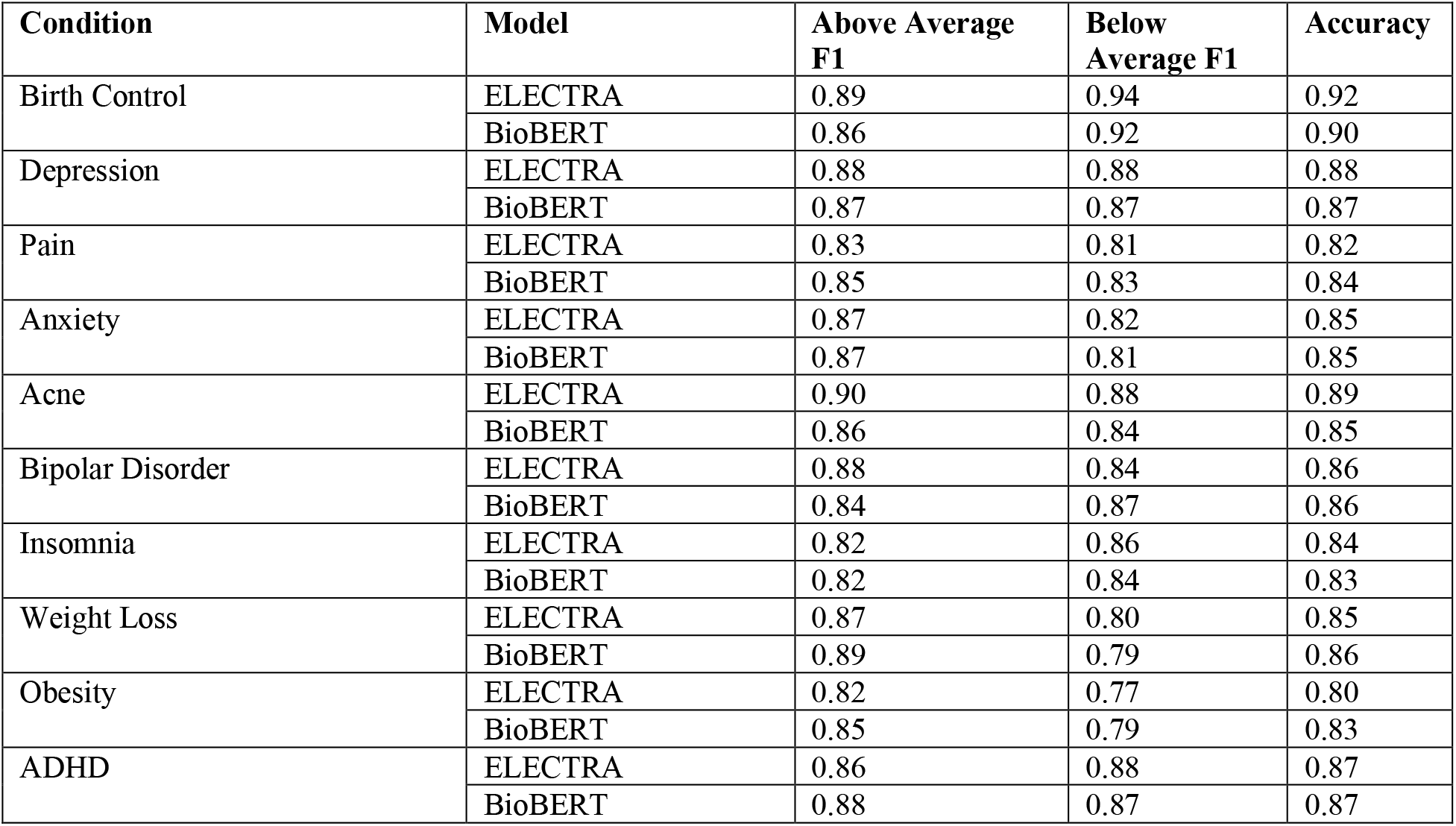
Results of condition-specific classifications for the top 10 conditions.

### Error Analysis

The results generated in Table 5 are produced by applying the Eli5 toolkit to the BioBERT trained model. There is a clear relationship between the words highlighted and the classification that was made by the model. Terms highlighted in green supports the classification generated by the model, while terms generated in red oppose the predictions. The shade of the color represents the level of importance at which a word contributes to the classification. Phrases related to side-effects were typically highlighted as *below-average* features such as being a “bit moody” or “sore”. The positive effects of the drug were highlighted as *above-average* features such as “my pain almost totally disappeared”. Specific highlighted terms by the ELi5 metric could potentially be subjected to an incorrect sentiment association. For example, the phrase “my cramps disappeared” in Table 5 for the false negative adverse event review was shown to support the prediction of a negative below average feature. However, this phrase would usually have a positive connotation associated with it unlike what is suggested by the model. In general, the major reason for a miss prediction by the model was primarily due to the presence of a mixture of positive and negative elements present in the text. This could have resulted from the presence of multiple medications, changes in the effectiveness of the medication over time, the extent of the medicinal effects, or treatment experience which could work in both sentimental directions to result in false positives or false. Adverse Events, in general, could result in false positive or false negatives depending on the extent to which the side-effect concerned someone. In addition, if a classification error occurred, a lower number of adverse events tended to be classified as false positive versus false negative. Overall, the interpretability of misclassifications, through the Eli5 tool kit revealed an important aspect of how the model used specific keywords.

**Table 5.**
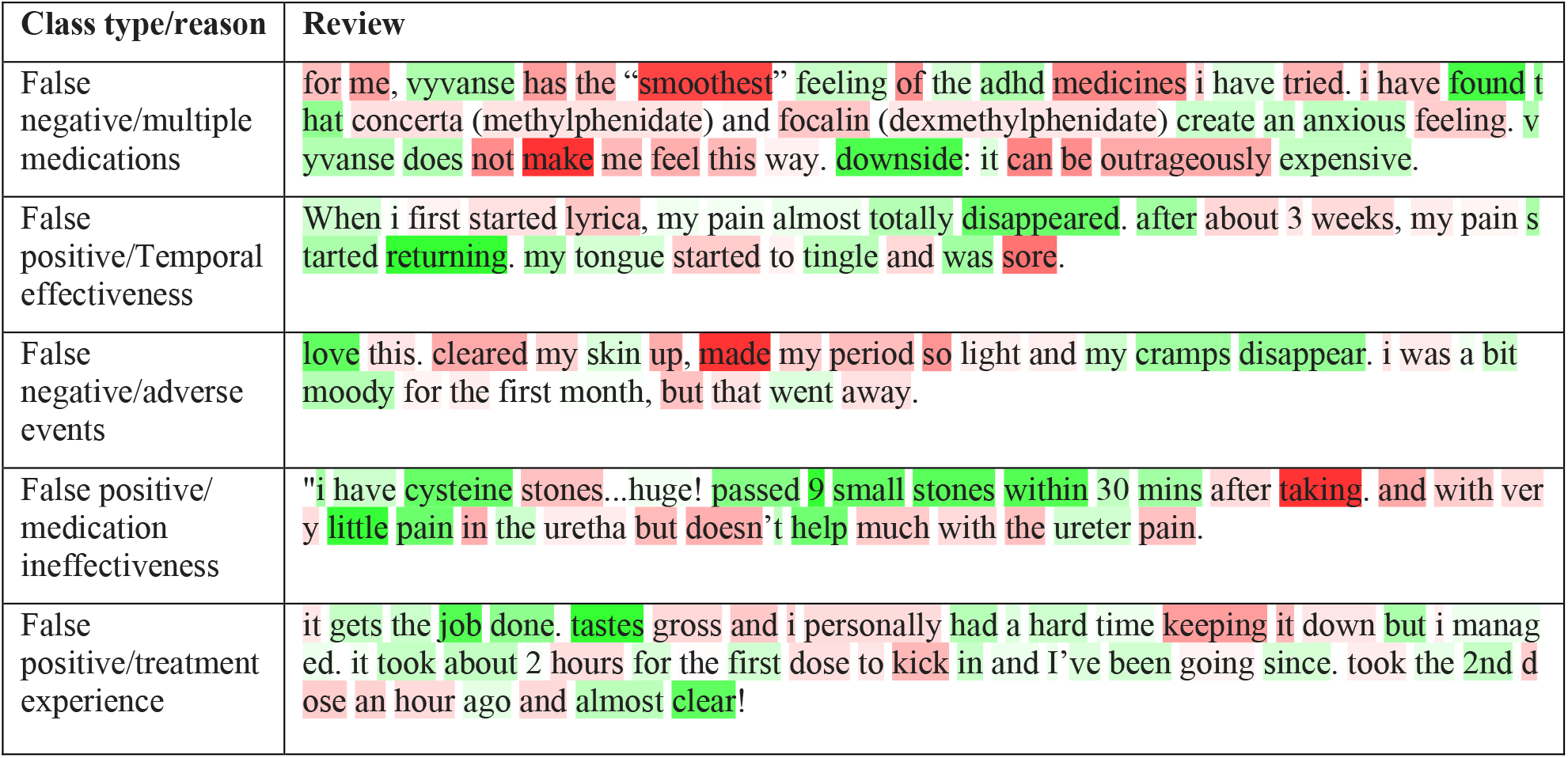
Examples of false positives and false negatives in BioBERT model with the important words highlighted in green (positively impacting the classification results) and red (negatively impacting the classification results) by the Eli5 toolkit.

Most semantic types were found to have a p-value < 0.05 for most of the class types based on the results shown in Table 6. The physiologic function semantic type for the BioBERT model and clinical drug name semantic type for both models were found to be insignificant. This suggests that both models tend not to heavily rely on the name of the clinical drug in predicting a score but could also be due to the lack of clinical drug names present in the user reviews. This idea is further supported based on the results of the ELi5 which shows many clinical drugs highlighted less impactful (lighter) to the classification than other terms in general. Based on results of the ELECTRA model, the average number of concepts of most sematic types (e.g., Sign or Symptom) in true negative instances is greater than that of true positive instances; and the average numbers of concepts of most sematic types in the false positive and false negative instances are between that of true positive and true negative instances

**Table 6.** The average mean number of semantic types reported for each class type based on the classification results of the overall BioBERT and ELECTRA models. P-values were generated using a 1-way ANOVA test for each condition using semantic type as the independent variable. Semantic types that differed from the most common pattern and were significant (α=0.05) are in bold.

| Model | Semantic Types | Sign or Symptom | Disease or Syndrome | Organism Function | Pathologic Function | Body Substance | Body Location or Region | Body Part, Organ, or Organ Component | Health Care Activity |
| --- | --- | --- | --- | --- | --- | --- | --- | --- | --- |
| ELECTRA | True Positive | 1.167 | 0.595 | 0.522 | 0.577 | 0.079 | 0.256 | 0.579 | 0.382 |
|  | True Negative | 2.317 | 0.925 | 0.867 | 0.879 | 0.153 | 0.435 | 0.944 | 0.534 |
|  | False Positive | 1.512 | 0.663 | 0.592 | 0.638 | 0.083 | 0.276 | 0.659 | 0.465 |
|  | False Negative | 1.733 | 0.750 | 0.778 | 0.716 | 0.137 | 0.366 | 0.814 | 0.516 |
|  | P-values | < 0.001 | < 0.001 | < 0.001 | < 0.001 | < 0.001 | < 0.001 | < 0.001 | < 0.001 |
| BIOBERT | True Positive | 1.374 | **0.697** | **0.623** | **0.675** | 0.097 | **0.307** | 0.688 | 1.374 |
|  | True Negative | 1.993 | **0.784** | **0.734** | **0.752** | 0.130 | **0.369** | 0.802 | 1.993 |
|  | False Positive | 1.705 | **0.784** | **0.725** | **0.706** | 0.111 | **0.345** | 0.781 | 1.705 |
|  | False Negative | 1.806 | **0.733** | **0.747** | **0.698** | 0.123 | **0.329** | 0.784 | 1.806 |
|  | P-values | < 0.001 | < **0.001** | < **0.001** | < **0.001** | < 0.001 | < **0.001** | < 0.001 | < 0.001 |

However, the BioBERT model has some more significant deviation from the most common class distribution in the ELECTRA model. Significant deviation from this class type distribution in the BioBERT model occur for Disease or Syndrome, Organism function, Pathologic function, Body region or Location.

All the implementation code and more examples of the misclassifications can be found in the GitHub repository.^32^

## Discussion

In this study, we built multiple classification models to classify drug review rating using the review text. Afterward, the Eli5 toolkit was applied to explain the models’ classification by highlighting the words that positively or negatively impacted the classification result. Informed by these experiments, it was clear that the consumers’ online drug reviews contain a vast quantity of information related to the sentiment expressed by the user. Transformer-based models have the potential to serve as a methodology to discriminate between overly positive and truly positive scores. Overall, this research outlined a potential process of identifying consumer drug review bias. This was consistent with other studies which found that subjective effects are often distorted in rating systems.^33^

When comparing different transformer-based models, the BioBERT and ELECTRA models outperformed the other models when the same amount of information was present. One possible reason for BioBERT to outperform other models is that BioBERT model was pre-trained with biomedical texts which were topically related to the drug reviews.^34^ According to the error analysis, ELECTRA model followed the pattern that the average numbers of concepts of most semantic types per post in true negative posts were greater than those in true positives; and those of false positive and false negative instances falling between those of true cases. This is further cemented by the fact that the ELi5 toolkit highlighted these terms as more important contributors in general. The BioBERT model tends to evade this classification for certain semantic types as previously stated in the error analysis. BioBERT trained with biomedical text allowed it to find more intricate relationships among the terms, allowing it to reach a better prediction accuracy. However, BioBERT and ELECTRA did significantly follow a similar pattern for the semantic types such as Sign or Symptom, Body substance, Body part organ or Organ component, and Health care activity. As it is clear that both models relied significantly on these factors to decern the sentiment of a user review, it is likely that user also weighted these factors higher than other semantic types when deciding their rating of the drugs. As such, higher number of possible adverse events (sign or symptoms), the need for more possible medical interventions (healthcare activities), and more reference to bodily fluids and organs (body substances and body part organ or organ component) tend to result in a lower rating.

Overly positive and negative scores given by the users could be detected by this model. Overly positive tend to suggest that the review given by the user does not reflect the positive rating that the user gave and conversely for overly negative scores. Disparities between user reviews and ratings may signify a knowledge gap for the score criteria or a certain level of subjectivity of the scoring.

Transformer models provide an automated, fast, and economic system to classify the sentiment of reviews from individuals for specific medications. Furthermore, transformer models’ capability to generate a suggestion of a score solely based on user reviews can be utilized as a point of comparison to user-generated reviews. In a clinical study, this discrepancy could potentially contribute towards advancing a conversation with the reviewer to further investigate the cause for such variations. In addition, ELi5 is an easy tool to understand what noteworthy terms contribute to the model, and potentially reviewers relied on providing more clarity on the logic behind the score. The identification of significant term contributors through the ELi5 metrics could hint at factors such as adverse events that are important in post-market drug surveillance. The binary classification approach of BioBERT and other transformer models could aid in potentially finding negative drug reviews in data that lacks a numeric score. This filtration of reviews delivers a vital step to simplify the process in the identification of adverse events, side effects, and possible medical interactions. A fast-paced system sentiment score prediction attests to its impact in analyzing large social media drug datasets providing a manageable tool to separate reviews into separate classes. The classified social media data can then be adopted for different purposes such as topic modeling by sentiment types.

### Limitations and Future Work

Although this model was able to successfully classify reviews in a binary system, the ability for large class identification is still unknown and warrants further investigation. One important issue with many transformer models was the issue of over-fitting.^35^ In addition, many transformer models such as XLNet are computationally expensive which may result in a long training time. Additional research will concentrate on the utilization of transformer models on non-scored-based social media data. In addition, another area of focus could be to expand this model for multi-class identification as this may be more advantageous in the determination of highly negative reviews.

## Conclusions

This study presents the construction of transformer-based models for the classification of drug reviews from drugs.com. The most successful model in this project was the BioBERT model with the highest F1 score. Overall, the transformer models outperformed the traditional machine learning models using bag-of-words features. These binary transformer models tended to be effective at decerning highly optimistic reviews from reviews that contain a mixture of positive and negative feedback.

## Data Availability

The dataset can be downloaded from https://archive.ics.uci.edu/ml/datasets/Drug+Review+Dataset+%28Drugs.com%29.

## Acknowledgements

This study was partially supported by the National Institute on Aging (NIA) of the National Institutes of Health (NIH) under Award Number R21AG061431; and in part by Florida State University-University of Florida Clinical and Translational Science Award funded by National Center for Advancing Translational Sciences under Award Number UL1TR001427. The first author would like to thank eHealth Lab at FSU and the Undergraduate Research Opportunity Program at Florida State University for the mentorship and guidance.

## References

1. Califf RM. Characteristics of clinical trials registered in clinicaltrials.gov, 2007-2010. JAMA;307(17):1838.

2. Farmer KC. Methods for measuring and monitoring medication regimen adherence in clinical trials and clinical practice. Clinical Therapeutic.1999;21(6):1074–90.

3. He Z, Tang X, Yang X, Guo Y, George TJ, Charness N, et al. Clinical trial generalizability assessment in the big data era: a review. Clin Transl Sci. 2020;13(4):675–84.

4. Mills EJ, Seely D, Rachlis B, Griffith L, Wu P, Wilson K, et al. Barriers to participation in clinical trials of cancer: a meta-analysis and systematic review of patient-reported factors. The Lancet Oncology. 2006;7(2):141–8.

5. Crombie I. The role of record linkage in post-marketing drug surveillance. British Journal of Clinical Pharmacology. 1986;22(S1):77S–82S.

6. Shimabukuro TT, Nguyen M, Martin D, DeStefano F. Safety monitoring in the vaccine adverse event reporting system (vaers). Vaccine. 2015;33(36):4398–405.

7. O’ Donovan B, Rodgers RM, Cox AR, Krska J. Making medicines safer: analysis of patient reports to the uk’s yellow card scheme. expert opinion on drug safety. 2019;18(12):1237–43.

8. Yom-Tov E, Gabrilovich E. Postmarket drug surveillance without trial costs: discovery of adverse drug reactions through large-scale analysis of web search queries. J Med Internet Res. 2013;15(6):e124.

9. Dey S, Luo H, Fokoue A, Hu J, Zhang P. Predicting adverse drug reactions through interpretable deep learning framework. BMC Bioinformatics. 2018;19(S21):476.

10. Ahmad F, Abbasi A, Kitchens B, Adjeroh DA, Zeng D. Deep learning for adverse event detection from web search. IEEE Trans Knowl Data Eng. 2020;1–1.

11. Moro PL, Arana J, Cano M, Lewis P, Shimabukuro TT. Deaths reported to the vaccine adverse event reporting system, united states, 1997–2013. Plotkin SA, editor. Clin Infect Dis. 2015;61(6):980–7.

12. Dandala B, Joopudi V, Devarakonda M. Adverse drug events detection in clinical notes by jointly modeling entities and relations using neural networks. Drug Saf. 2019;42(1):135–46.

13. Kulldorff M, Davis RL, Kolczak† M, Lewis E, Lieu T, Platt R. A maximized sequential probability ratio test for drug and vaccine safety surveillance. Sequential Analysis. 2011;30(1):58–78.

14. Du J, Xiang Y, Sankaranarayanapillai M, Zhang M, Wang J, Si Y, et al. Extracting postmarketing adverse events from safety reports in the vaccine adverse event reporting system (vaers) using deep learning. Journal of the American Medical Informatics Association. 2021;

15. Dinh T, Chakraborty G. Detecting side effects and evaluating the effectiveness of drugs from customers’ online reviews using text analytics, sentiment analysis, and machine learning models. sas-global-forum-proceedings. 2020;1–23.

16. Hino A, Imai R. Ranking and rating: neglected biases in factor analysis of postmaterialist values. International Journal of Public Opinion Research. 2019;31(2):368–81.

17. Tanabe P, Buschmann M. A prospective study of ed pain management practices and the patient’s perspective. Journal of Emergency Nursing. 1999;25(3):171–7.

18. Adusumalli S, Lee H, Hoi Q, Koo S-L, Tan IB, Ng PC. Assessment of web-based consumer reviews as a resource for drug performance. J Med Internet Res. 2015;17(8):e211.

19. Lewis DD. Challenges in machine learning for text classification. In: Proceedings of the ninth annual conference on Computational learning theory - COLT ‘96. Desenzano del Garda,, Italy: ACM Press. 1996;1–ff.

20. UCI. drug review dataset (drugs.com) data set. Available from: https://archive.ics.uci.edu/ml/datasets/Drug+Review+Dataset+%28Drugs.com%29

21. Sairamvinay Vijayaraghavan, Debraj Basu. Sentiment analysis in drug reviews using supervised machine learning algorithms. Available from: https://arxiv.org/abs/2003.11643

22. Gräßer F, Kallumadi S, Malberg H, Zaunseder S. Aspect-based sentiment analysis of drug reviews applying cross-domain and cross-data learning. In: Proceedings of the 2018 International Conference on Digital Health. Lyon France: ACM. 2018;121–5.

23. Devlin J, Chang M-W, Lee K, Toutanova K. BERT: pre-training of deep bidirectional transformers for language understanding. arxiv: 181004805. 2019;

24. Lee J, Yoon W, Kim S, Kim D, Kim S, So CH, et al. BioBERT: a pre-trained biomedical language representation model for biomedical text mining. Wren J, editor. Bioinformatics. 2019;

25. Lan Z, Chen M, Goodman S, Gimpel K, Sharma P, Soricut R. ALBERT: a lite bert for self-supervised learning of language representations. arxiv: 190911942. 2020;

26. Liu Y, Ott M, Goyal N, Du J, Joshi M, Chen D, et al. RoBERTa: a robustly optimized bert pretraining approach. arxiv: 190711692. 2019;

27. Yang Z, Dai Z, Yang Y, Carbonell J, Salakhutdinov R, Le QV. XLNET: generalized autoregressive pretraining for language understanding. arxiv: 190608237. 2020;

28. Clark K, Luong M-T, Le QV, Manning CD. ELECTRA: pre-training text encoders as discriminators rather than generators. arxiv: 200310555. 2020;

29. Agarwal N, Das S. Interpretable machine learning tools: a survey. 2020 IEEE Symposium Series on Computational Intelligence (SSCI). Canberra, ACT, Australia: IEEE. 2020;1528–34.

30. Bodenreider O. The unified medical language system (umls): integrating biomedical terminology. Nucleic Acids Research. 2004;32(90001):267D –270.

31. Luca Soldaini, Nazli Goharian. QuickUMLS: a fast, unsupervised approach for medical concept extraction. 2016; Available from: http://medir2016.imag.fr/data/MEDIR_2016_paper_16.pdf

32. akhilfsu/Classifying-Drug-Ratings-Using-User-Reviews-with-Transformer-Based-Language-Models. Available from: https://github.com/akhilfsu/Classifying-Drug-Ratings-Using-User-Reviews-with-Transformer-Based-Language-Models

33. Abou Taam M, Rossard C, Cantaloube L, Bouscaren N, Roche G, Pochard L, et al. Analysis of patients’ narratives posted on social media websites on benfluorex’s (Mediator ^®^) withdrawal in France. J Clin Pharm Ther [Internet]. 2014;39(1):53–5.

34. Pipalia K, Bhadja R, Shukla M. Comparative analysis of different transformer based architectures used in sentiment analysis. In: 2020 9th International Conference System Modeling and Advancement in Research Trends (SMART). Moradabad, India: IEEE. 2020;411–5.

35. Komatsuzaki A. One epoch is all you need. arxiv: 190606669. 2019;

